# Workforce guidelines, burnout, work engagement and the intention to leave. A cohort study among Dutch Emergency Physicians

**DOI:** 10.1101/2025.11.17.25340438

**Authors:** Annemieke E. Boendermaker, Gideon H.P. Latten, Jelle T. Prins, Kiki M.J.M.H. Lombarts, Paul L.P. Brand

## Abstract

**Background and importance:** Emergency medicine (EM) physicians work in a dynamic and challenging environment, with high risk of burnout. The prevalence of work engagement, burnout and the intention to leave among Dutch EM physicians have not been studied before. This study aims to evaluate the adherence to the DSEP workforce recommendations, and the association between these recommendations and signs of burnout, work engagement and the intention to leave the profession.

**Methods:** Cross-sectional survey-based cohort study among all Dutch EM physicians. Burnout and work engagement were assessed using the Dutch version of the Maslach Burnout Inventory and the Utrecht Work Engagement Scale.

**Main results:** Overall, 295 EM physicians participated (response rate 56%, 68% female, median age 39 years). Fifty-five respondents (18.5%) met the criteria for burnout. Compared to other Dutch healthcare providers, EM physicians scored significantly higher on depersonalization and lower on personal accomplishment. The burnout rate among Dutch EM physicians compared favourably to those reported in international literature. Work engagement was higher compared to studies among other Dutch medical specialists. Intention to leave the profession was high with 22-29%. Adherence to the workforce recommendations varied widely between hospitals.

**Conclusions:** Almost a fifth of Dutch EM physicians experience burnout and adherence to workforce recommendations varied widely. Work engagement amongst Dutch EM physicians was relatively high. Intention to leave was high. To ensure a sustainable future in which EM physicians are protected from burnout, supported in their work, and where disproportionate job turnover is minimized, it is crucial these results are met with decisive action. This study creates a foundation upon which future research can build and a repeating questionnaire is already planned for end 2025.

- **What is already known on this topic –** *In many fields in healthcare workforce guidelines are developed to improve physician wellness and carreer longevity. Burnout and job turnover in emergency medicine are high*.
- **What this study adds –** *This study is the first looking into burnout, work engagement and intention to leave among Dutch Emergency physicians. Furthermore it relates its workforce guideline directly to these outcomes, underlining its importance and providing a baseline for future research and improvements*.
- **How this study might affect research, practice or policy –** *Further analysis as planned and better implementation of workforce guidelines could in the future help diminish burnout and job turnover and improve work engagement and physician wellness*.

## INTRODUCTION

Emergency medicine (EM) physicians work in a fast-paced, unpredictable setting, where stressful situations are common. To maintain wellbeing, work engagement and quality of life, it is important to invest in a sustainable work environment, aimed at preventing burnout and improving career longevity. In 2015, the Dutch Society of Emergency Physicians (DSEP/NVSHA) published its workforce guideline for durable employability and scheduling, stipulating a set of core recommendations and conditions to aspire to.^1^ These recommendations are based upon extensive research and similar guidelines from the UK, USA and Australasia.^2–4^ Nevertheless, literature on the prevalence of burnout among Dutch EM physicians, and international literature on the effects of adherence to workforce recommendations is lacking.

Burnout is recognised worldwide as a syndrome related to a person’s profession. It consists of three domains, which are separately measured: emotional exhaustion, depersonalization and a sense of diminished personal accomplishment.^5–8^ Burnout in healthcare providers is associated with diminished mental and physical health, but also with a reduced quality of work, a rise in patient related safety incidents, an increase in the intention to leave the profession and a lasting incapacity for work.^9–11^

The opposite of burnout is considered to be work engagement; a positive, fulfilling, work-related state of mind in its own right that is characterised by three psychometrically distinct aspects: vigor, dedication, and absorption.^12–14^ Where burnout is mainly predicted by job demands and a lack of job resources, work engagement is exclusively predicted by available job resources.^11–14^ Work engagement fosters the intention to remain in the profession through a motivational process, stimulates extra-role behaviour, enhances overall wellbeing, and protects against patient related safety incidents. ^11–14^

The core recommendations for Dutch EM physicians outlined in the workforce guideline address factors presumed to influence burnout and work engagement, such as scheduling and contract conditions. However, these recommendations have not yet been studied as a set. In this cross-sectional cohort study, we aim to estimate the levels of burnout and work engagement experienced by Dutch EM physicians prior to the COVID-19 pandemic, assess adherence to the different workforce recommendations and examine the association between compliance with these recommendations and the wellness parameters of burnout, work engagement, and intention to leave.

## METHODS

### Design and setting

This cross-sectional survey-based cohort study was conducted among all 526 Dutch EM physicians (DSEP) at the time. From May 1^st^ to June 30^th^ 2019, all were invited via personal e-mail to complete an online questionnaire developed by a multidisciplinary research team of EM physicians, psychologists, epidemiologists, medical education researchers and human resource management specialists (Supplementary Online Content – Questionnaire_Dutch.docx). The questionnaire included internationally validated questions to assess burnout, work engagement and intention to leave the profession, and demographic characteristics, and work situation. After two weeks, nonresponders received a reminder to complete the questionnaire.

The study was approved by a regional Dutch ethics review board (RTPO-1046). All participants provided informed consent by checking the consent box at the start of the questionnaire. To further ensure data and privacy safety demographics were separated from the main dataset. Only the two main researchers had the key to link both data sets for analysis purposes.

### Variables and definitions

Demographics – Respondents were asked to provide data on age, gender, relationship status and family composition, year of graduation from medical school, and year of completion of postgraduate EM physician training.

Burnout – The Utrecht Burnout Scale (UBOS), the validated Dutch version of the Maslach Burnout Inventory, was used.^15^ Respondents reported their agreement with statements on the three separate domains of burnout - emotional exhaustion, depersonalization and sense of diminished personal accomplishment - using a 7-point Likert scale, ranging from “never” (1) to “always” (7). For the definition of ‘burn out’, we used the Brenninkmeijer EEplus1 definition, which requires the presence of emotional exhaustion and either high depersonalization or low personal accomplishment.^16^ When emotional exhaustion, high depersonalization and low personal accomplishment were all present, respondents fulfilled the definition of ‘burnout syndrome’ and were also considered to be burned out.^17^ In line with the UBOS manual, the 75^th^ percentile of the distribution of this composite variable was used as a cut-off for burnout. The scales were compared to the norm reference values of Dutch healthcare personnel presented in the UBOS manual.^15^

Work engagement – The 3-item version of the Utrecht Work Engagement Scale (UWES), which assesses vigor, dedication and absorption, was used.^18^ Each item was scored on a 5-point Likert scale from “never” (1) to “always” (5). Overall work engagement was defined and analysed as a composite variable of all three items and compared to the norm reference values provided by the UWES manual of work engagement scores of Dutch healthcare professionals.^19^

Work conditions – The work situation was assessed with a model derived from the Dutch Workforce Guideline for Durable Employability and Scheduling.^1^ This workforce guideline offers 12 recommendations for working conditions, which were translated to dichotomous questions regarding EM physicians’ working conditions. These included scheduled backward rotation, fixed cycle in rotation scheduling, fixed shift off, break time during shift, weekends per month, nightshift to team size, shift duration, contract size, additional scheduled time, presence of ED residents, team size to ED size, and contract type.

Intention to leave the profession^20–21^ – Intention to leave was assessed using a 4-point Likert scale, ranging from never (1) to multiple times (4). EM physicians were asked whether, given the choice, they would choose the same profession again ‘knowing what they know now’, and whether they saw themselves working as an EM physician in 10 years’ time?” The choice of a 10-year time horizon was based on the expected young age of the respondent group, given the relative novelty of EM physician as a profession in the Netherlands.

### Analysis and statistics

To assess the profession’s representativeness of the sample population, we compared respondents’ gender and age to those of all registered EM physician members of the DSEP. Descriptive statistics were used to describe the prevalence of burnout and adherence to the 12 workforce recommendations (Figure 1). We assessed differences in demographic and working conditions using variables between respondents with or without a positive value on the burnout composite score using chi-square tests, Fisher exact tests or Mann-Whitney-U tests, as appropriate. Two sided p values <0.05 were considered statistically significant. Cronbach’s alpha was used to measure the internal consistency of construct variables.

To assess correlation between burnout, work engagement, intention-to-leave and the workforce recommendations, we computed Spearman rank (r_s_), rank biserial and phi correlation coefficients.

All analyses were performed using IBM SPSS Statistics for Windows, version 27 (IBM Corp., Armonk, N.Y., USA).

## RESULTS

### Demographics

In total, 295 (56%) EM physicians completed the questionnaire, of which 200 (67.8%) were female. Median age was 39 (IQR 35-44) years and respondents had been working as an EM physician for a median of 6 (IQR 3-10) years. Age and sex were comparable between respondents and the entire cohort of DSEP members (Supplementary Table 1 – E_table_1.docx).

### Burnout symptoms and work engagement

Burnout, defined as emotional exhaustion plus high depersonalization and/or low personal accomplishment, was observed in 55 (18.6%) respondents (Table 1). High emotional exhaustion was present in 27.1%, and low personal accomplishment in 21.6%. Consistent with the UBOS manual, depersonalisation was reported as a male score at 25.0% and female score of 28.0%.

**Table 1.** Demographics and burnout. Numbers are median (IQR) or n (%)

| Table 1 – Demographics and burnout |  |  |  |  |
| --- | --- | --- | --- | --- |
| Demographics | Total (n=295) | No burnout (n=240) | Burnout (n=55) | p |
| Sex - female | 200 (67.8%) | 164 (68.3%) | 36 (65.5%) | 0.75 |
| Age – years | 39 (35-44) | 40 (35-43) | 41 (36-45) | 0.22 |
| Working as EM physicians – years | 6 (3-10) | 6 (3-10) | 6 (3-9) | 0.98 |
| With partner | 247 (83.7) | 209 (87.1%) | 38 (69.1%) | 0.002 |
| With children | 196 (66.4) | 159 (66.3%) | 37 (67.3%) | 0.89 |

On work engagement, 29 (16.6%) respondents scored low to extremely low (i.e. the 25^th^ groups’ percentile) and 97 (32.9%) respondents scored high to extremely high (i.e. the groups’ 75^th^ percentile).

When compared to the UBOS and UWES reference scores, the EM physicians in this study did not score significantly different on emotional exhaustion scores (1.86 vs. 1.78, p=0.31), but they did score significantly lower on personal accomplishment (4.12 vs. 4.22, p=0.03) and overall work engagement (3.48 vs. 3.74, p<0.01) (Table 2).^15;19^

**Table 2.** Burnout and work engagement scales compared to norm reference values of Dutch healthcare personnel. Burnout scales in reference to the validated norm reference values of Dutch healthcare personnel presented in the UBOS manual and Dutch physicians of the UWES manual.^15;19^ * The UBOS manual applies gender-specific reference values to depersonalization scale due to consistently significant differences between male and female subjects, explained in terms of prevailing gender roles.

| Burnout scales | Emergency physicians (n=295) |  | Reference group (n=10,552) |  | p | 95% CI |
| --- | --- | --- | --- | --- | --- | --- |
|  | Mean | SD | Mean | SD |  |  |
| Emotional exhaustion | 1.86 | 1.15 | 1.78 | 1.21 | 0.31 | -0.07 - 0.23 |
| Depersonalization |  |  |  |  |  |  |
| - Women | 1.21 | 0.78 | 1.14 | 0.78 | 0.12 | -0.02 - 0.16 |
| - Men | 1.45 | 1.11 | 1.27 | 0.84 | <0.01 | 0.08 - 0.28 |
| Diminished personal accomplishment | 4.12 | 0.70 | 4.22 | 0.78 | 0.03 | -0.19 - -0.01 |

Table 2. Burnout scales in reference to the validated norm reference values of Dutch healthcare personnel presented in the UBOS manual and Dutch physicians of the UWES manual.<sup>15,19</sup> \* The UBOS manual applies gender-specific reference values to depersonalization scale due to consistently ...
| Work engagement scales | Emergency physicians (n=295) |  | Reference group (n=655) |  | p | 95% CI |
| --- | --- | --- | --- | --- | --- | --- |
|  | Mean | SD | Mean | SD |  |  |
| Overall work engagement | 3.48 | 0.65 | 3.10 | 1.92 | <0.01 | -0.61 - -0.16 |
| Subscales |  |  |  |  |  |  |
| - Vigor | 3.39 | 0.79 | 3.04 | 0.92 | <0.01 | -0.47 - -0.23 |
| - Dedication | 3.78 | 0.81 | 3.29 | 1.04 | <0.01 | -0.62 - -0.36 |
| - Absorption | 3.26 | 0.89 | 2.96 | 1.92 | 0.011 | -0.53 - -0.07 |

### Intention to leave

Based on a Cronbach’s alpha of 0.62 - considered a weak correlation - the three questions on intention to leave were assessed separately instead of taken together. Almost a third of respondents (29%) answered having considered leaving the profession several or multiple times. The consideration to leave the profession was significantly more common amongst respondents with burnout (n=55, 36%) than those without (n=240, 10%, p<0.001). Sixty-eight respondents (23%) did not see themselves still working as EM physicians in 10 years’ time. This was also more common in physicians with burnout (49%) than those without (17%, p<0.001). Physician age did not explain this difference (39.7 ± 5.7 vs 41.0 ± 5.8 years, 95% CI of difference −0.3 to 2.9 years, p = 0.103). Similarly, 23% of respondents reported not choosing EM as a career again (49% in burned out vs 18% in not burned out physicians, p <0.001).

Table 3 shows the odds ratios of burnout and work engagement on the intention to leave questions.

**Table 3.** Odds ratios of burnout and work engagement on the intention to leave questions.

|  | Burnout |  |  | Work engagement |  |  |
| --- | --- | --- | --- | --- | --- | --- |
|  | Odds ratio | 95% CI | p | Odds ratio | 95% CI | p |
| Positive future | 0.21 | 0.11 – 0.39 | <.001 | 3.3 | 2.1 – 5.2 | <.001 |
| Choosing EM again | 0.22 | 0.1 – 0.4 | <.001 | 4.2 | 2.6 – 6.6 | <.001 |
| Seldom to never considering to quit | 0.23 | 0.1 – 0.2 | <.001 | 3.5 | 2.3 – 5.4 | <.001 |

### Working conditions

Table 4 shows the adherence of respondents’ EDs to the 12 dichotomized workforce recommendations and their association with burnout. There were differences in the adherence to the recommendations. The mean of followed recommendations was 6.6 with a range of 1 to 11. The most often followed recommendation (99.0%) was to work with designated ED residents, while the least commonly followed recommendation (8.3%) was the use of a fixed cycle in rotation scheduling.

**Table 4.** Adherence in the respondents’ ED to the 12 workforce recommendations and association with burnout. Numbers are n (%)

| Recommendation | Answers | N (%) | Burnout | No burnout | p |
| --- | --- | --- | --- | --- | --- |
| Scheduled backwards rotation | 0-1x/month | 220 (76.9%) | 40 (18.2%) | 180 (81.8%) | 0.78 |
|  | >1/month | 66 (23.1%) | 13 (19.7%) | 53 (80.3%) |  |
| Fixed cycle in rotation scheduling | Yes | 24 (8.3%) | 7 (29.2%) | 17 (70.8%) | 0.17 |
|  | No | 265 (91.7%) | 47 (17.7%) | 218 (82.3%) |  |
| Fixed shift off per week | Yes | 165 (56.3%) | 26 (15.8%) | 139 (84.2%) | 0.17 |
|  | No | 128 (43.7%) | 29 (22.7%) | 99 (77.3%) |  |
| No break during shift | Never/sometimes | 298 (58.8%) | 8 (9.5%) | 76 (90.5%) | 0.01 |
|  | Often/always | 209 (41.2%) | 47 (20.9%) | 162 (77.5%) |  |
| Working full weekends per month | 1 in 3-5 weekends | 68 (23.2%) | 8 (11.8%) | 60 (88.2%) | 0.09 |
|  | 1 in 1-2 weekends | 225 (76.8%) | 47 (20.9%) | 178 (79.1%) |  |
| Appropriate team size for nightshifts | Yes | 182 (66.2%) | 29 (15.9%) | 153 (84.1%) | 0.25 |
|  | No (too small) | 93 (33.8%) | 20 (21.5%) | 73 (78.5%) |  |
| Shift duration 9 hours | Yes | 180 (61.0%) | 26 (14.4%) | 154 (85.6%) | 0.02 |
|  | No | 115 (39.0%) | 29 (25.2%) | 86 (74.8%) |  |
| Contract size per week | 32-38 hours | 208 (70.3%) | 36 (17.3%) | 172 (82.7%) | 0.36 |
|  | <32 or >38 hours | 88 (29.7%) | 19 (21.8%) | 68 (78.2%) |  |
| Additional scheduled time | ≥15% | 63 (23.2%) | 8 (12.7%) | 55 (87.3%) | 0.18 |
|  | <15% | 209 (76.8%) | 42 (20.1%) | 167 (79.9%) |  |
| Working with designated ED residents | Yes | 292 (99.0%) | 54 (18.5%) | 238 (81.5%) | 0.51 |
|  | No | 3 (1.0%) | 1 (33.3%) | 2 (66.7%) |  |
| Team size in relation to ED size | Sufficient | 32 (11.9%) | 5 (15.6%) | 27 (84.4%) | 0.82 |
|  | Insufficient | 238 (88.1%) | 41 (17.2%) | 197 (82.8%) |  |
| Contract type | Permanent | 258 (87.5%) | 43 (16.7%) | 215 (83.3%) | 0.02 |
|  | Temporary | 37 (12.5%) | 12 (32.4%) | 25 (67.7%) |  |

### Associations

Three workforce recommendations were associated with an increased risk of burnout when not adhered to: the presence of breaks during shifts (p=0.01), a shift duration of 9 hours (p=0.02), and a permanent contract (p=0.02) (Table 4).

Work engagement was associated with working 1 in 3-4 weekends or less (p=0.03). Intention to leave the profession was associated with not adhering to the following recommendations: working 1 in 3-4 weekends or less (p=0.01) and working in a team large enough to match the ED size (p=0.05).

## DISCUSSION

### Main findings and interpretation

In this study, we examined adherence to Dutch workforce guidelines for EM physicians and investigated the association between compliance with these recommendations and levels of burnout, work engagement and intention to leave the profession. Adherence to workforce recommendations varied with a mean of only 6.6 followed: the recommendation to work with designated ED residents was followed most frequently (99.0%), while the use of a fixed cycle in rotation scheduling was adhered to least (8.3%). Not adhering to three specific workforce recommendations was significantly associated with burnout: the absence of breaks during shifts, a shift duration of less or more than 9 hours, and a temporary contract. Work engagement was associated with working 1 in 3-4 weekends or less (p=0.03). Intention to leave the profession was associated with not adhering to the following recommendations: working 1 in 3-4 weekends or less (p=0.01) and working in a team large enough to match for the ED size (p=0.05).

Comparing the prevalence of burnout in our population of Dutch EM physicians (18.6%) with that reported in other studies using the Maslach burnout inventory proved less straightforward than expected, mainly due to the use of different measurements, scales and different cut-off values.^22^ The study of Meynaar et al from 2015 showed a comparable way of calculating burnout, but reported a surprising low prevalence of 4.4% among 272 Dutch intensivists (41% response rate), which the authors hypothesised was due to the low workload in Dutch ICUs.^23^ A previous meta-analaysis, including over 1250 EM physicians from 12 countries, showed high levels of emotional exhaustion and depersonalization (both around 40%) and low personal accomplishment in 35% of EM physicians.^23^ While these numbers are higher than those observed in our study, they are not directly comparable due to the use of various different cut-offs. Despite our relatively favorable results, we must recognize that 1 in 5 Dutch EM physicians in our study displayed signs of burnout. Given the potential negative consequences for both physicians and patients, this prevalence is concerning and underscores the need for intervention. ^9–11^ Adhering to the workforce recommendations presents one avenue for improvement, as table 4 suggests.

Since our study is the first to investigate the adherence to the Dutch workforce guidelines, direct comparisons with other studies is challenging. Nevertheless, the workforce recommendations are based on previously published research and similar strategies for preventing burnout have been reported by others.^2–4,13,25–27^ Time during a shift to take a break, recover and have designated time to eat are all proven to be beneficial for physician wellness.^27–31^ Working 9 hours provides overlap between colleagues to safely transfer care of patients and to end the shift before concentration and energy levels diminish.^31^ The job securiy that comes with a permanent contract besides the impact it has on team building might explain this recommendation to be assosiated with burnout.

### Strengths and limitations

Our study is the first to investigate the prevalence of burnout, work engagement and intention to leave among Dutch EM physicians, and the adherence to the Dutch workforce recommendations, establishing a baseline for future efforts to promote a sustainable EM workforce. It was the first nationwide study of its kind and the response rate was 56% which we consider high in comparison with similar survey studies among physicians. The use of validated instruments, such as the Dutch version of the Maslach Burnout Inventory (UBOS) and the Utrecht Work Engagement Scales (UWES) allows for future comparisons. However, this study did not account for the potential effects of the COVID-19 pandemic. Therefore, repeating this research in the current Dutch EM environment is recommended.

## Data Availability

Data collection was performed by Triple i Human Capital research group. https://www.3ihc.nl/. The DSEP has the proprietary right to the data, with access limited to members of the research group.

## Conclusion

We found that almost 1 in 5 Dutch EM physicians exhibited signs of burnout, just prior to the COVID-19 pandemic. On the opposite, levels of work engagement were higher compared to other Dutch medical specialists. Adherence to the DSEP workforce recommendations promises to be an effective strategy to prevent burnout, and job turnover and promote work engagement. Three recommendations in particular deserve to be addressed: the presence of breaks during shifts, a shift duration of 9 hours, and the availability of a permanent contract. Repeating this study to obtain updated prevalence rates is planned for the end of 2025, concurrent with the publication of the revised Workforce guideline. The results of this current study were used in its revision and provide the basis of a longitudinal analysis. The overall aim is to create better work conditions and terms of employment for health care professionals.

